# Clinical utility of measuring CD4^+^ T follicular cells in patients with immune dysregulation

**DOI:** 10.1101/2023.06.06.23291032

**Authors:** Brenna LaBere, Alan A. Nguyen, Saddiq B. Habiballah, Megan Elkins, Juliet Imperial, Betty Li, Sridevi Devana, Suraj Timilsina, Spencer B. Stubbs, Jill Joerger, Janet Chou, Craig D. Platt

## Abstract

Mechanistic studies of autoimmune disorders have identified circulating T follicular helper (cTfh) cells as drivers of autoimmunity. However, the quantification of cTfh cells is not yet used in clinical practice due to the lack of age-stratified normal ranges and the unknown sensitivity and specificity of this test for autoimmunity. We enrolled 238 healthy participants and 130 patients with common and rare disorders of autoimmunity or autoinflammation. Patients with infections, active malignancy, or any history of transplantation were excluded. In 238 healthy controls, median cTfh percentages (range 4.8% – 6.2%) were comparable among age groups, sexes, races, and ethnicities, apart from a significantly lower percentages in children less than 1 year of age (median 2.1%, CI: 0.4% – 6.8, *p<*0.0001). Among 130 patients with over 40 immune regulatory disorders, a cTfh percentage exceeding 12% had 88% sensitivity and 94% specificity for differentiating disorders with adaptive immune cell dysregulation from those with predominantly innate cell defects. This threshold had a sensitivity of 86% and specificity of 100% for active autoimmunity and normalized with effective treatment. cTfh percentages exceeding 12% distinguish autoimmunity from autoinflammation, thereby differentiating two endotypes of immune dysregulation with overlapping symptoms and different therapies.

## 1. Introduction

Autoimmune disorders affect approximately 4.5% of the world’s population and continue to rise in complexity and prevalence. Common autoimmune disorders are classified by clinical, laboratory, and/or genetic criteria. However, a subset of patients has disorders of multifaceted autoimmunity leading to organ dysfunction without fulfilling clinical criteria that define other autoimmune disorders [1]. These disorders are a notable diagnostic and therapeutic challenge due to their clinical heterogeneity, rarity, and multisystemic involvement, underscored by the development of rare disease action plans and autoimmune alliances in 23 countries [2]. Next-generation DNA sequencing has identified over 344 genetic causes of autoimmunity or autoinflammation [3], but most patients lack a genetic diagnosis. More targeted therapeutics may be used if autoimmune diseases can be distinguished from those with predominantly autoinflammatory mechanisms [4,5]. However, the clinical symptoms associated with autoimmunity and autoinflammation are similar [6]. Current diagnostic approaches, which include the measurement of autoantibodies, histologic analysis of immune cell infiltrates, and genetic testing, are diagnostic in only a minority of cases. Furthermore, laboratory and radiologic imaging may not detect worsening immune dysregulation until end-organ dysfunction occurs.

Studies of clinically and genetically define autoimmune disorders have identified a role for T follicular helper cells in causing autoimmunity. These cells are a subtype of CD4^+^ T cells in secondary lymphoid organs that generate pathogenic autoantibodies as well as protective antibodies [7]. T follicular helper cells express CXCR5^+^, a chemokine receptor that enables migration to B cell follicles [8], and PD-1, a marker of T cell activation [9]. T and B cell interactions in secondary lymphoid organs generate circulating T follicular helper (cTfh) cells, which exit the lymph node through efferent lymphatic vessels into the blood [10]. An elevated percentage of cTfh cells is associated with autoimmunity, evidenced by cTfh expansion in common autoimmune diseases, including SLE [11], rheumatoid arthritis [12], and type I diabetes mellitus [13], as well as in rare monogenic disorders [14–17]. Small cohort studies of select disorders, including SLE, LRBA deficiency, CTLA4 haploinsufficiency, and Evans syndrome have indicated that a clinical response to effective therapy is often accompanied by reduction of cTfh percentage [11,14,18–20]. Despite the low cost and feasibility of measuring cTfh cells by flow cytometry, this test is not yet used in clinical practice. There are no published studies establishing the normal range for cTfh percentage through childhood development, and the sensitivity and specificity of cTfh percentage for identifying active autoimmunity is unknown.

Here, we establish the normal range for cTfh percentage using 234 immunologically healthy individuals ranging from neonates to young adults. We demonstrate the utility of measuring cTfh percentage in characterizing disorders of multifaceted autoimmunity, thereby addressing a clinical gap in the disorders lacking unified clinical criteria.

## 2. Materials and Methods

### 2.1. Study sample

Patients were recruited at Boston Children’s Hospital. The study was approved by the Institutional Review Board of Boston Children’s Hospital. Patients within the healthy category were stratified by age as well as by race/ethnicity, as self-designated in the medical chart. Patients designated as “Black” were those who identified as non-Hispanic Black and/or non-Hispanic African American. Those designated as “Hispanic” identified as Hispanic and/or Latino, and Spanish, Portuguese, or Puerto Rican. Patients designated as “other” included a self-identification of “other” race, with ethnicities listed as Chinese, Indian, Arab, or other. Patients within the “unknown” category were labeled as “unknown/not reported.” Those designated as “White” included patients who self-identified as White, Caucasian, French Canadian, French, European, Irish, and German.

Study participant characteristics are further described in the results. Disease activity was abstracted from the medical record by the study’s clinical immunologists prior to measurement of cTfh cells to minimize investigator bias. “Active disease” was defined by the presence of clinical symptoms, examination findings, or laboratory, radiologic, or other diagnostic abnormalities indicative of active disease, as reported by medical providers in the electronic medical record. “Quiescent” or “controlled” disease was defined by an absence of physical complaints or significant changes in laboratory, radiologic, other diagnostic testing in the patients’ medical records.

### 2.2. Flow cytometry

Surface staining for CD3 (BD, Cat# 340948), CD4 (BD, Cat# 347413), CD8 (BD, Cat# 655111), PD-1 (BioLegend, Cat# 329907), and CXCR5 (Invitrogen, Cat# 12-9185-42) was performed on whole blood by incubating with monoclonal antibodies for 15 minutes, followed by lysis in BD FACS Lysing Solution (diluted 1:10 in MilliQ water). Cells were centrifuged at 1500 rpm for 5 minutes, followed by resuspension in 200 uL phosphate buffer saline. Data were collected with a FACSCanto cytometer (BD Biosciences) and analyzed with FlowJo software (TreeStar, Ashland, Ore). PD-1^+^CXCR5^+^ cells were defined as a percentage of the total CD3^+^CD4^+^ T cell population (**Supplementary Figure 1**), with a stop gate of 10,000 CD3^+^ events. We found that the staining of cTfh cells was comparable on blood samples stored for 24 hours at 4°C compared to room temperature (**Supplementary Figure 2**). cTfh percentages were highly reproducible between the three flow cytometers used in the analysis (**Supplementary Figure 3**), indicating minimal intermachine variability.

### 2.3. Statistical analysis

The reference ranges and confidence intervals for cTfh cells were determined using the non-parametric methods from Reed et al., as recommended by the Clinical and Laboratory Standards Institute [21]. In this approach, the upper and lower limits for the 2.5 percentile and 97.5 percentile were determined based on a ranked cTfh levels from 210 immunologically healthy individuals over 1 year of age. We used receiver operator characteristic curve (ROC) analysis to determine sensitivity and specificity of cTfh for differentiating (1) autoimmunity from autoinflammation and (2) quiescent from active autoimmune disease. ROC analysis was performed using IBM^®^SPSS^®^ Statistics (v. 28) and GraphPad Prism (v. 9.4.0). Comparison between groups was carried out with Mann-Whitney test, Dunn’s multiple comparison test, and multi-comparison Kruskal-Wallis test, as indicated. Differences were considered significant at a *p* value of less than 0.05, with adjustment for multiple comparisons as indicated in figure legends.

## 3. Results

### 3.1. Patient characteristics

Patients were delineated into different categories as shown in **Figure 1**. The 364 participants in this study ranged from 3 days to 35 years of age (**Table 1**). The healthy control cohort consisted of 234 patients younger than 35 years of age who had no evidence of infection at the time of sample collection and no history of immune or autoimmune disorders, malignancy, hematopoietic stem cell transplantation, or treatment with immunomodulatory therapies. All controls had normal absolute lymphocyte counts at the time of the assay. We compared the normal ranges to cTfh percentages in patients with rheumatologic disorders, including 13 patients with SLE, a disorder associated with expansion of cTfh cells [22,23], and 31 patients with juvenile idiopathic arthritis (JIA) (including oligo-, mono-, and polyarticular JIA, systemic JIA, and psoriatic arthritis), in whom cTfh percentage has previously been reported as normal [24,25]. We then quantified cTfh cell percentage in 55 patients with multifaceted autoimmunity and 31 patients with autoinflammatory disorders. Primary immune regulatory disorders caused by inborn errors of immunity were based on criteria from the 2022 Update on the Classification from the International Union of Immunological Societies Expert Committee [3]. All patients categorized as having multifaceted autoimmunity met one of the following criteria 1) two organ systems involved, 2) one organ system involved with a monogenic primary immune regulatory disorder diagnosis, or one organ system if presentation is frequently associated with a monogenic diagnosis (e.g. Evan’s syndrome); 45% of patients in this cohort had concomitant immunodeficiency. A genetic diagnosis was known in 47% of those with multifaceted autoimmunity and 26% of those with autoinflammatory disorders, with over 90% of this cohort having undergone extensive genetic evaluation (**Supplementary Table 1**). The clinical features of the patients with genetically undefined multifaceted autoimmunity are detailed in **Table 2.** No patients had a history of solid organ or hematopoietic stem cell transplantation, active treatment of malignancy, or infection at the time of study enrollment. Each disease category contained patients with active and controlled disease.

**Figure 1:**
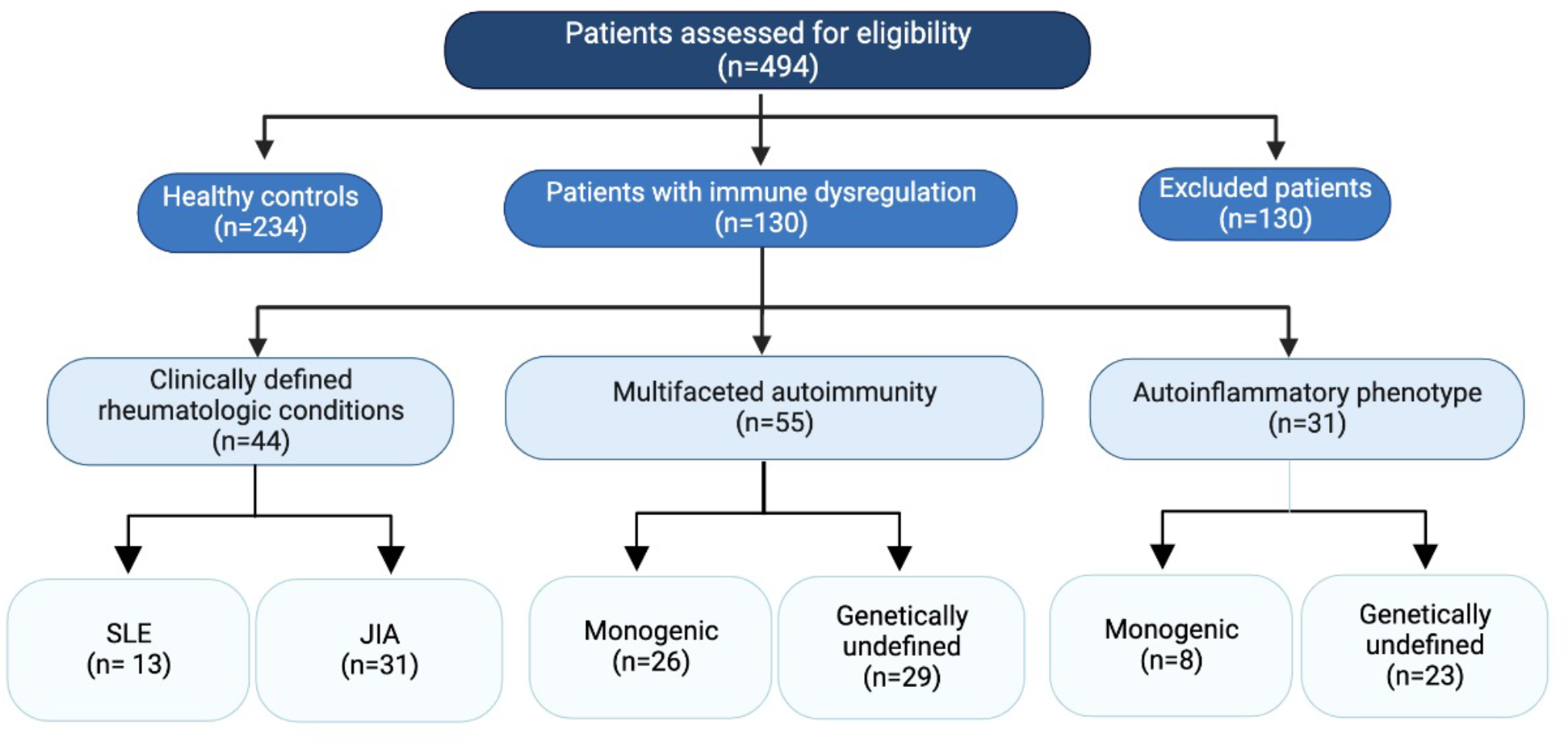
Study design. Of all patients assessed for eligibility, 364 were classified as being healthy controls versus having some type of immune dysregulation. Those with immune dysregulation were further divided based on phenotype/diagnosis, as delineated in the schematic. JIA: juvenile idiopathic arthritis; SLE: systemic lupus erythematosus.

**Table 1:**
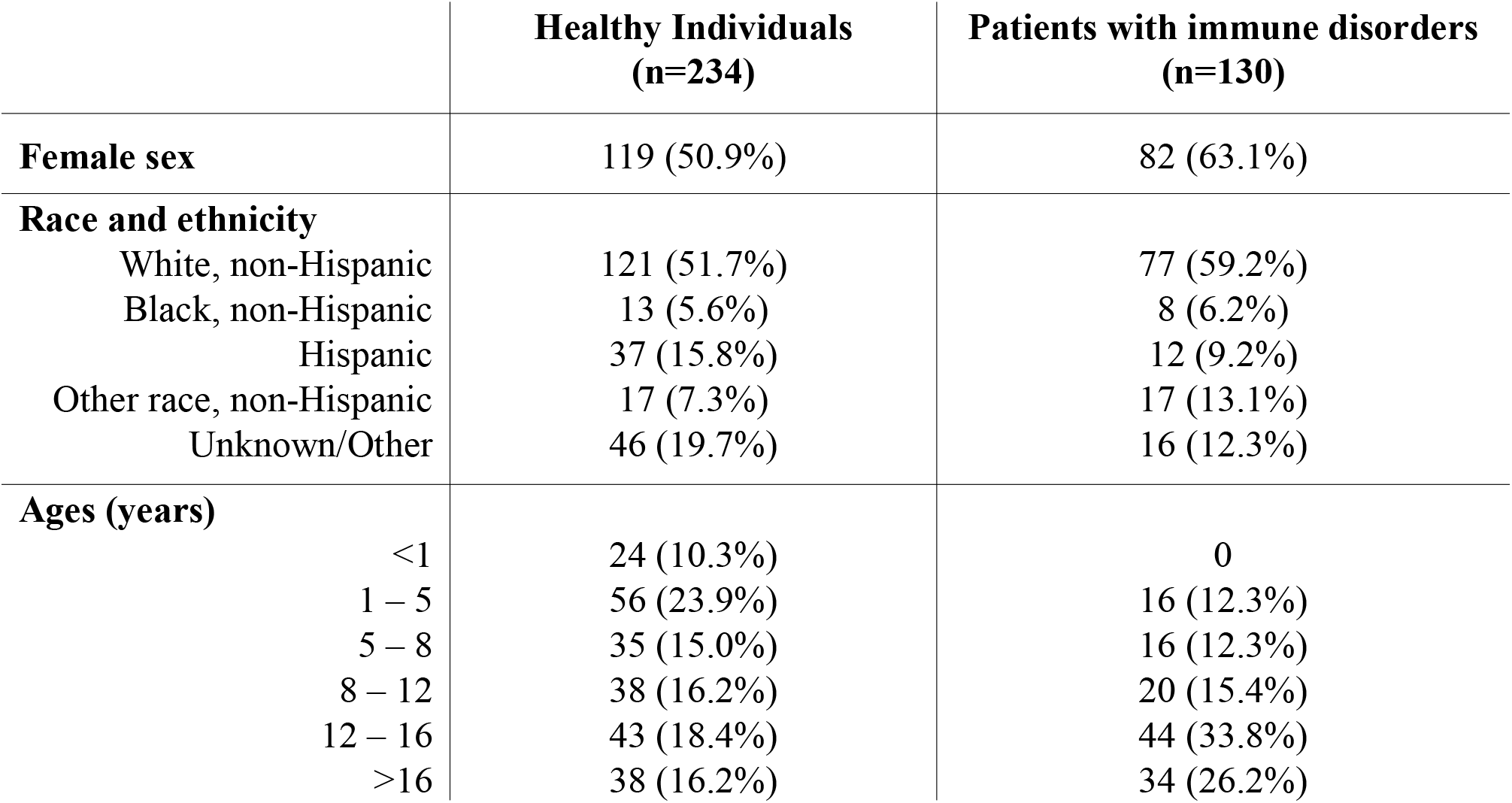
Patient characteristics.

|  | Healthy Individuals<br>(n=234) | Patients with immune disorders<br>(n=130) |
| --- | --- | --- |
| Female sex | 119 (50.9%) | 82 (63.1%) |
| Race and ethnicity |  |  |
| White, non-Hispanic | 121 (51.7%) | 77 (59.2%) |
| Black, non-Hispanic | 13 (5.6%) | 8 (6.2%) |
| Hispanic | 37 (15.8%) | 12 (9.2%) |
| Other race, non-Hispanic | 17 (7.3%) | 17 (13.1%) |
| Unknown/Other | 46 (19.7%) | 16 (12.3%) |
| Ages (years) |  |  |
| <1 | 24 (10.3%) | 0 |
| 1 – 5 | 56 (23.9%) | 16 (12.3%) |
| 5 – 8 | 35 (15.0%) | 16 (12.3%) |
| 8 – 12 | 38 (16.2%) | 20 (15.4%) |
| 12 – 16 | 43 (18.4%) | 44 (33.8%) |
| >16 | 38 (16.2%) | 34 (26.2%) |

**Table 2:**
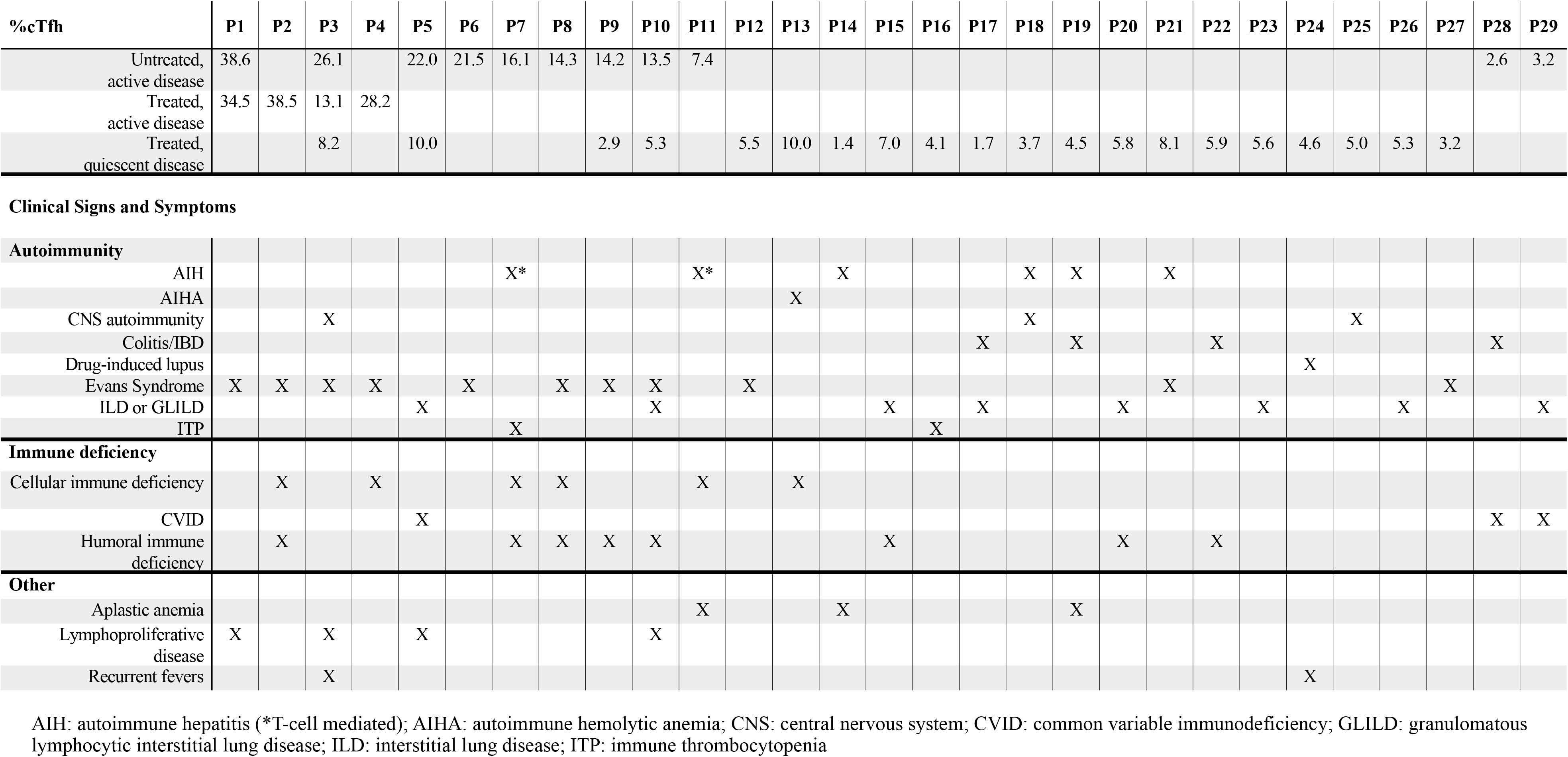
Characteristics of patients with genetically undefined multifaceted autoimmunity.

### 3.2. cTfh percentages in healthy individuals

CXCR5^+^PD1^+^ cTfh cells were measured as a percentage of CD4^+^ T cells. Within healthy individuals, cTfh percentages were comparable between males and females and among races and ethnicities (**Figures 2A and 2B**). Patients younger than one year of age had significantly lower cTfh percentages than those over one year of age, but these values were comparable among all individuals over one year of age (**Figures 2C and 2D**). The median cTfh percentage in 210 healthy individuals older than one year of age was 5.2% (90% confidence interval for 2.5 percentile: 1.3 – 2.3%; 90% confidence interval for 97.5 percentile: 9.4 – 11.3%). We then compared this reference range to cTfh percentage in patients with JIA and SLE, as prior studies have characterized cTfh levels in these disorders [11,18,24–28]. cTfh percentage in patients with JIA were comparable to that of healthy individuals, as has been previously published [24,25]. This is concordant with the known contribution of innate cells and T peripheral helper cells, both which of which are distinct from Tfh cells, to JIA pathogenesis [28,29]. In contrast, the median cTfh percentage was significantly increased in patients with active SLE (**Figure 3A**), reflecting the known contributions of cTfh cells to the development of autoantibodies in this disease [22,23]. cTfh percentage was normal in patients with well-controlled SLE, also concordant with previously published studies. This was the case even in patients with chronic disease and end organ involvement, if on effective therapy, demonstrating that cTfh percentage correlates with disease activity. This measurement thus representing a useful tool for assessing response to therapies [30].

**Figure 2:**
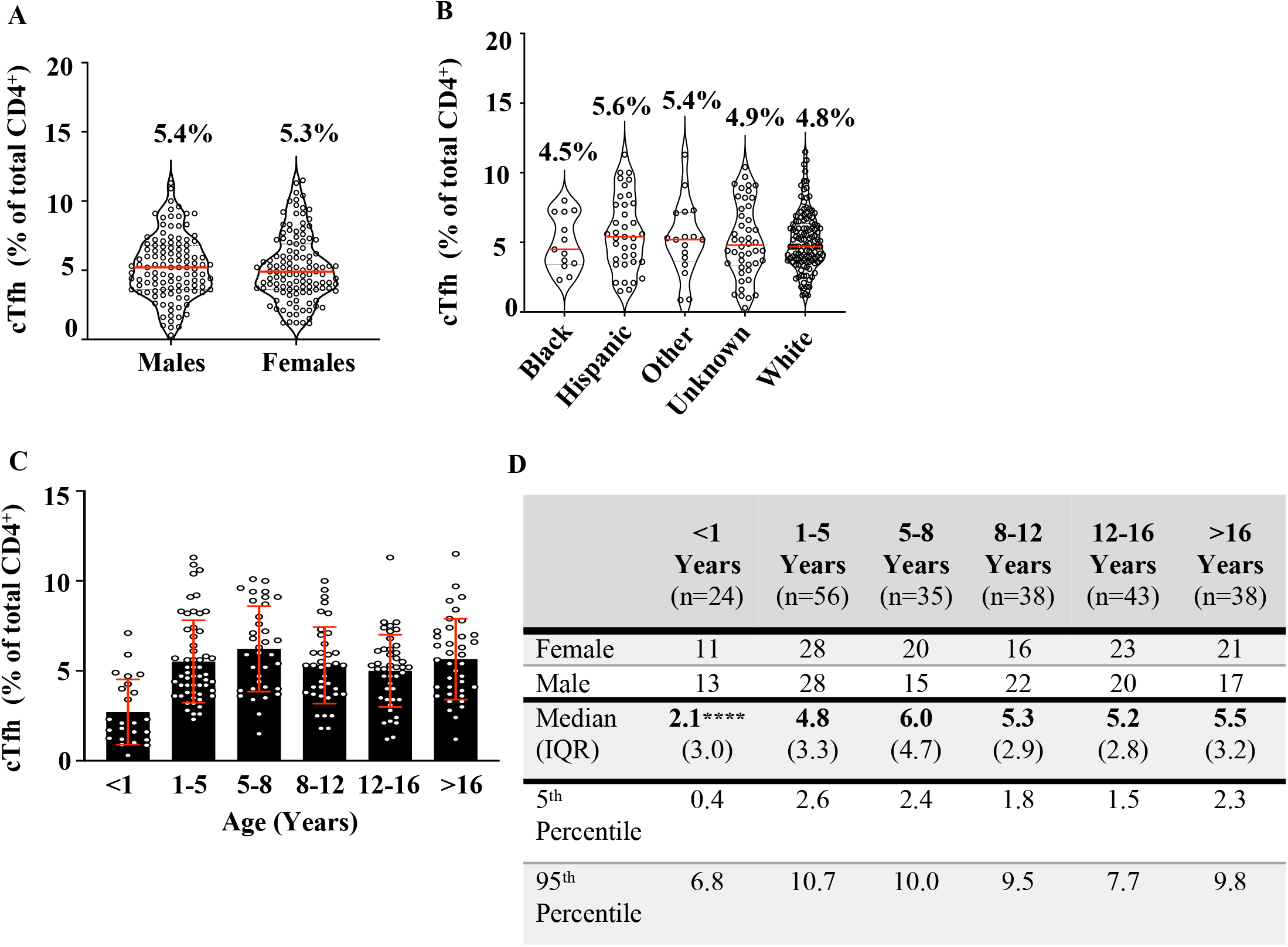
Percentages of CD4^+^CXCR5^+^PD1^+^ cTfh cells (expressed as a percentage of total CD4^+^) in healthy control patients grouped by sex (A), self-reported race/ethnicity (B) and age (C&D), with median cTfh percentages reported for each group within the figure for sex and self-reported race/ethnicity. *p*=ns by Mann-Whitney test for sex and multiple-comparison Kruskal-Wallis test for race/ethnicity. \*\*\*\**p*<0.0001 for age comparison, as determined by multi-comparison Kruskal-Wallis test.

**Figure 3:**
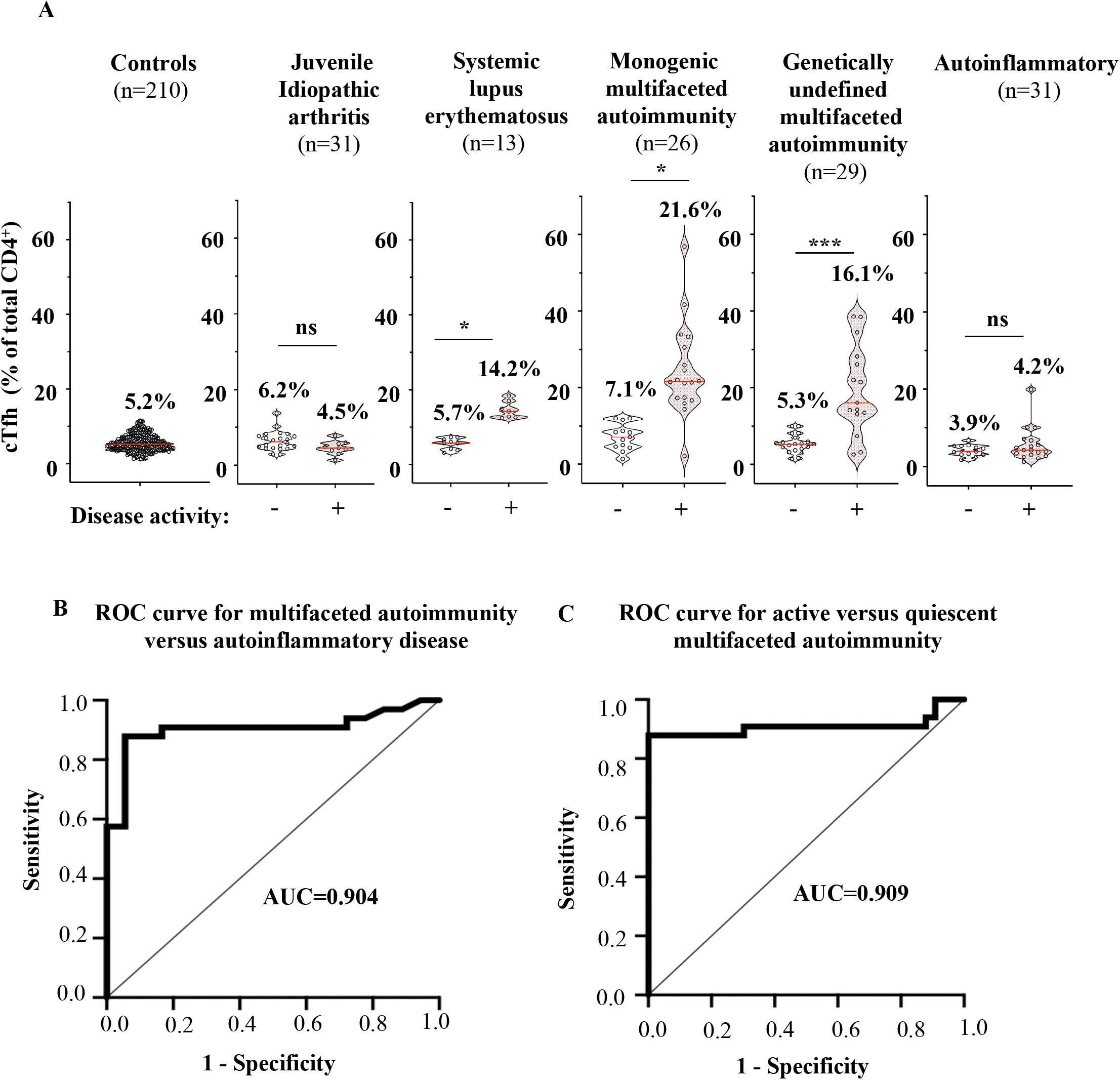
Percentages of CD4^+^CXCR5^+^PD1^+^ cTfh cells in patients with immune dysregulation with quiescent (-) and active (+) disease, with *n* indicating the total number of patients within each category. The median cTfh percentage is indicated for each group (A). Receiver operator curves (ROC) demonstrating sensitivity and specificity of cTfh frequencies in distinguishing patients with multifaceted autoimmunity from those with autoinflammatory disease (B), and in distinguishing patients with active versus quiescent multifaceted autoimmunity (C). AUC, area under the curve; JIA: juvenile idiopathic arthritis; MA: multifaceted autoimmunity; SLE: systemic lupus erythematosus. *\*p*<0.05, \*\*\**p*<0.001 by Dunn’s multiple comparisons test.

### 3.3. Applications of cTfh percentages in patients with multifaceted autoimmunity

Unlike SLE and JIA, most patients with multifaceted autoimmunity lack clinical classification and diagnostic criteria [31]. Although over 340 monogenic causes of primary immune regulatory disorders have been identified, the majority of patients lack a genetic diagnosis. The diagnosis of these diseases is further complicated by their clinical heterogeneity, which encompasses cytopenias, interstitial lung disease, gastrointestinal inflammation, and neuroinflammation, some of which can overlap with the symptoms of autoinflammatory disorders, which are mediated by the innate immune system [5]. It was unknown if patients with genetically-defined multifaceted autoimmunity have similar levels of cTfh to those lacking a genetic diagnosis after extensive genetic testing (i.e. whole exome sequencing). Compared to controls, median cTfh percentages were increased in patients with active disease and multifaceted autoimmunity, irrespective of the underlying genetic diagnosis (**Figure 3A**). In contrast, the cTfh percentages were normal in patients with well-controlled disease (**Figure 3A**). We performed receiver operating characteristic (ROC) curve analysis to determine how well cTfh expansion discriminates between autoimmunity and autoinflammation. Among patients with active disease, the ROC curve indicated that a cTfh cell expansion has high diagnostic performance in distinguishing diseases with evidence of adaptive immune dysfunction from autoinflammatory disorders, with an area under the curve (AUC) of 0.904 (**Figure 3B**). The threshold of cTfh cell expansion that optimizes sensitivity and specificity in discriminating multifaceted autoimmunity from autoinflammatory diseases is 12% (sensitivity 88% and specificity of 94%).

We next determined how well cTfh cell expansion correlates with disease activity in patients with multifaceted autoimmunity. This is clinically relevant since other approaches, including imaging, organ biopsies, endoscopies, and colonoscopies, are more invasive, expensive, and/or logistically challenging than the measurement of cTfh cell percentage. Among patients with multifaceted autoimmunity, an ROC curve analysis indicates that an expansion of cTfh cells had high diagnostic performance in identifying patients with active autoimmunity, with an AUC of 0.909. A cTfh percentage greater than 12% identified patients with active autoimmunity, with a sensitivity of 86% and specificity of 100% (**Figure 3C)**.

To illustrate practical applications of these findings, we present two clinical vignettes. Our study included one patient with Activated PI3K Delta syndrome (APDS), a congenital immune disorder with diverse clinical symptoms including autoimmunity, immunodeficiency, and susceptibility to malignancy. At a routine follow-up visit, the patient was completely asymptomatic, with stable anemia and lymphopenia, normal liver and renal function tests, and no abnormalities noted on physical examination. However, the patient was found to have an elevated cTfh percentage at 21.4%, suggesting a need for additional evaluation to identify the source of active disease. Pulmonary function testing revealed severe restrictive lung disease with forced expiratory volume less than 40% predicted, and computerized tomography identified interstitial lung disease. This case thus illustrates the utility of measuring cTfh levels to identify patients with poorly controlled autoimmunity.

In counterpoint, Patient 5 illustrates how declining cTfh levels can reflect clinical improvement. This patient has common variable immune deficiency, granulomatous lymphocytic interstitial lung disease, lymphadenopathy, and hepatosplenomegaly, but no genetic diagnosis. The patient’s cTfh percentage was 22% before initiation of any immunomodulation. After starting abatacept, an immunomodulatory agent that reduces T cell co-stimulation, the cTfh level declined to 10% in parallel with radiologically confirmed reductions in lymphadenopathy and pulmonary nodules. Similar reductions occurred in patients on whom we had longitudinal measurements of cTfh levels (**Table 2**). These cases illustrate how longitudinal measurements of cTfh levels may be useful in monitoring disease activity in genetically undefined disorders, thus broadening the clinical applicability of cTfh beyond specific rare diseases or clinically well-defined disorders [32–36].

## 4. Discussion

Despite the uses of cTfh percentages in autoimmunity research, barriers have prevented adoption in clinical practice. Our study addresses the specific gaps preventing utilization of cTfh cells in the evaluation of patients. First, we establish reference ranges for cTfh percentages using a cohort of 234 healthy children and adults, which will enable future clinical studies of cTfh percentages in other types of immune disorders. Except for infants, cTfh percentages are similar among healthy children and young adults, thereby increasing the utility of this assay. We show that children under one year of age have significantly lower median cTfh percentages (median 2.1%, CI: 0.4% – 6.8%) compared to that of children in older age groups (median cTfh levels ranging from 4.8% to 6.0%, **Figure 2D**). The reduced cTfh percentages in children under one year of age is relevant to the susceptibility of infants to viral illnesses, such as respiratory syncytial virus, rhinovirus, and influenza [37,38]. A recent study showed that native CD4^+^ T cells from neonatal mice secrete increased levels of the cytokine IL-2, an important factor for establishing peripheral tolerance that also reduces the generation of Tfh cells in early life [33]. Our findings thus provide a human corollary to these mechanistic studies in mice, since previously published studies of cTfh percentages in children are few and have focused on populations with active infections, such as malaria, human immunodeficiency virus, and measles [40–42].

Our findings indicate that cTfh percentage distinguishes between multifaceted autoimmunity and autoinflammatory disorders, two disease types with overlapping clinical features [43,44]. The normal cTfh percentages in patients with active autoinflammatory disease likely reflect the contributions of innate immunity to these disorders. Clinical tests for distinguishing autoimmunity from autoinflammation include genetic testing and cytokine panels, which can be costly or inaccessible [4,45,46]. The measurement of many cytokines is affected by sample processing time, storage, and number of freeze-thaw cycles. In contrast, the cTfh assay is an inexpensive test requiring less than 30 minutes of hands-on technician time, and is reproducible when performed on whole blood that has been *ex vivo* for up to 24 hours. We estimate that the clinical costs for cTfh measurement is approximately $150, based on costs of similar commercial flow cytometry tests with a comparable number of markers and complexity, compared to $2500 for whole exome sequencing and $450 for a 13-plex cytokine panel. Thus, cTfh percentage expands the diagnostic tool set available for differentiating active autoimmune disease from autoinflammation, a distinction that aids the diagnosis and treatment of these diseases.

Additionally, cTfh percentage greater than 12% correlated with disease activity in patients with multifaceted autoimmunity, regardless of genetic diagnosis. Our study thus increases the applicability of measuring cTfh percentage beyond what has been previously published in patients with rare monogenic diseases and Evans syndrome [14,18,20,49–54]. We show that an elevated cTfh percentage is not a marker of any specific genetic diagnosis, but instead correlates with active disease in those with multifaceted autoimmunity. Additional studies are necessary to determine if quantification of cTfh cells can be used to titrate immunosuppressive medications or if patients with elevated cTfh percentages are more likely to respond to particular therapies.

### 4.1. Limitations

Our study has limitations. This was a single center study. We excluded patients with clinical signs and symptoms of infections, given the confounding effect of infections on increasing cTfh cell percentages [55–57]. Additional studies are needed to examine the impact of infections on cTfh percentages in patients with immune dysregulation. As our study aimed to first determine the utility of measuring total cTfh percentages in a patient population with a wide range of immune disorders, we did not investigate the further sub-phenotypes of cTfh cells in these patients. Subtypes include the cTfh1 subtype characterized by expression of IFN-γ, the cTfh2 and cTfh13 subtypes characterized by distinct expression profiles of IL-4, IL-13, and IL-5, and the cTfh17 subtype characterized by expression of IL-17 [50,53,54,58]. Skewing of cTfh subtypes has been identified in small studies of patients with immune dysregulation [50,53,54,58,59]. We also acknowledge that some disorders may have aspects of both autoimmunity and autoinflammation, and that additional studies of cTfh levels are needed in these types of mixed disorders.

## 5. Conclusions

Although the increasing availability of genomic sequencing has accelerated the diagnosis of monogenic disorders, there remains a need for immunophenotypic and functional assays that correlate with disease activity. Enumeration of cTfh percentages is measured by flow cytometry with inexpensive, rapid surface staining of whole blood. This study indicates that increased levels of cTfh cells are a sensitive and specific indicator of active disease in patients with multifaceted autoimmunity, and that these levels normalize with effective treatment. By stratifying disease activity, the quantification of cTfh cells complements advances in genomics for patients with diverse autoimmune endotypes.

## Data Availability

All data produced in the present study are available upon reasonable request to the authors.

## Abbreviations

AIH: autoimmune hepatitis
AIHA: autoimmune hemolytic anemia
cTfh: circulating T follicular helper cell
CNS: central nervous system
CVID: common variable immune deficiency
GLILD: granulomatous lymphocytic interstitial lung disease
ILD: interstitial lung disease
IRD: immune regulatory disorder
ITP: immune thrombocytopenia
JIA: juvenile idiopathic arthritis
sJIA: systemic juvenile idiopathic arthritis
SLE: systemic lupus erythematosus
Tfh: T follicular helper cell

## Figure Legends

**Supplementary Figure 1:** Representative flow cytometry analysis used to define CD4+PD1^+^CXCR5^+^ cTfh cells and flow cytometry analysis for a healthy control patient versus a patient with genetically undefined active immune dysregulation (A). Sample stability; cTfh% measured at indicated timepoints after storage at 4 degrees Celsius or room temperature. *n*=5 samples. \*\**p*=0.0020 by 2-tailed *t* test. Comparison of laboratory flow cytometers; the percentage of cTfh cells was determined in 11 peripheral blood samples on each of 3 lab instruments and the results were plotted on linear regression plots. R^2^ values are noted (C).

**Supplementary Figure 2:** Sample stability. cTfh% measured at indicated timepoints after storage at 4 degrees Celsius or room temperature. n=5 samples. \*\**p*=0.0020 by 2-tailed *t* test.

**Supplementary Figure 3:** Comparison of laboratory flow cytometers. The percentage of Tfh cells was determined in 11 peripheral blood samples on each of 3 lab instruments and the results were plotted on linear regression plots. R^2^ values are noted.

