## Supplemental Table 1 for "Clinical utility of measuring CD4^+^ T follicular cells in patients with immune dysregulation"

**Supplementary Table 1: Diagnoses within immune dysregulation categories**

| **Category**. For diseases with descriptive or eponymous names, the affected gene is listed in parentheses. | **Number of Patients (%)**  **n=130** |
| --- | --- |
| **Genetically defined primary immune regulatory disorders** | **26** (20.0%) |
| LRBA haploinsufficiency* | 6 |
| CTLA4 haploinsufficiency* | 6 |
| STAT3 gain-of-function | 2 |
| Autoimmune lymphoproliferative syndrome (*FAS*) | 1 |
| Activated PI3K-delta syndrome (*PIK3CD)* | 1 |
| Hermansky-Pudlak syndrome (*AP3B1)* | 1 |
| Ataxia telangiectasia (*ATM*) | 1 |
| X-linked agammaglobulinemia (*BTK*) | 1 |
| Immune dysregulation, polyendocrinopathy, enteropathy, X-linked syndrome (*FOXP3*) | 1 |
| Kabuki Syndrome due to a pathogenic *KMT2D* variant | 1 |
| NFKB1 haploinsufficiency | 1 |
| RIPK1 deficiency | 1 |
| SOCS1 haploinsufficiency | 1 |
| TNFRSF13B deficiency with autoimmunity | 1 |
| Wiskott-Aldrich syndrome (*WAS*) | 1 |
| **Genetically undefined primary immune regulatory disorders** | **29** (22.3%) |
| **Systemic lupus erythematosus** | **13** (10.0%) |
| **Inflammatory arthritis** | **31** (23.8%) |
| Juvenile idiopathic arthritis, seronegative | 11 |
| Juvenile idiopathic arthritis, seropositive | 8 |
| Psoriatic arthritis | 8 |
| Systemic juvenile idiopathic arthritis | 4 |
| **Monogenic autoinflammatory disorders** | **8** (6.2%) |
| Cryopyrin-associated periodic syndrome (*NLRP3*) | 2 |
| Aicardi Goutières syndrome (*RNASE2HA* and *RNASEH2B*) | 2 |
| A20 haploinsufficiency (*TNFAIP3*) | 1 |
| ADA2 deficiency | 1 |
| Neonatal-onset cytopenia with autoinflammation, rash, and hemophagocytosis (*CDC42*) | 1 |
| STING-associated vasculopathy with onset in infancy | 1 |
| **Genetically undefined autoinflammatory disorders** | **23** (17.7%) |
| Recurrent fever syndromes, not specified | 13 |
| Chronic recurrent multifocal osteomyelitis | 5 |
| Periodic fever, aphthous stomatitis, pharyngitis, and adenitis | 3 |
| Bechet’s disease | 2 |

*The clinical and laboratory details of nine patients with CTLA4 haploinsufficiency or LRBA deficiency have been previously published^10^
