## Supplemental Figures 1, 2, and 3 for "Clinical utility of measuring CD4^+^ T follicular cells in patients with immune dysregulation"

### Slide 1
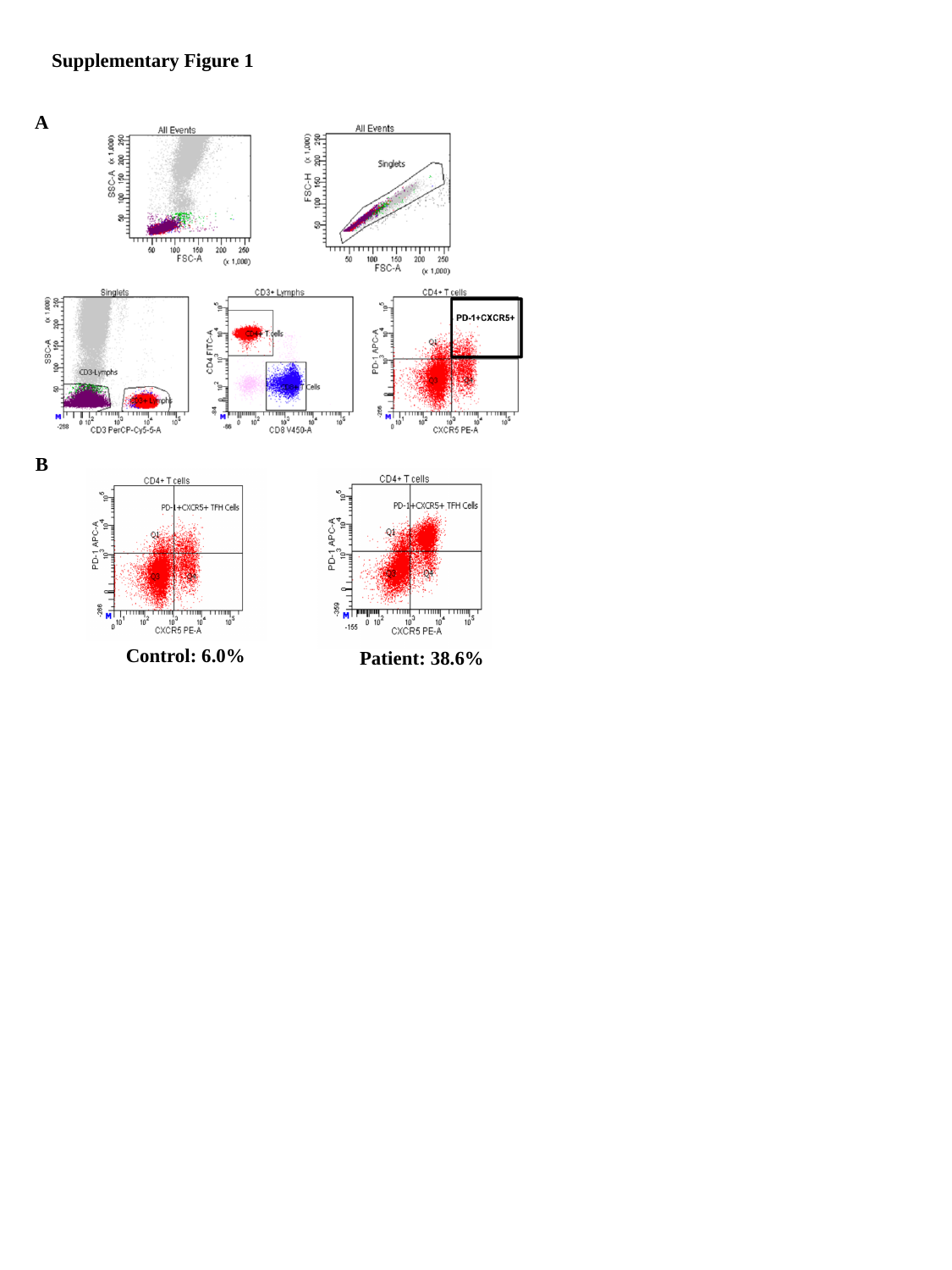

Supplementary Figure 1
A
B
Control: 6.0%
Patient: 38.6%

### Slide 2
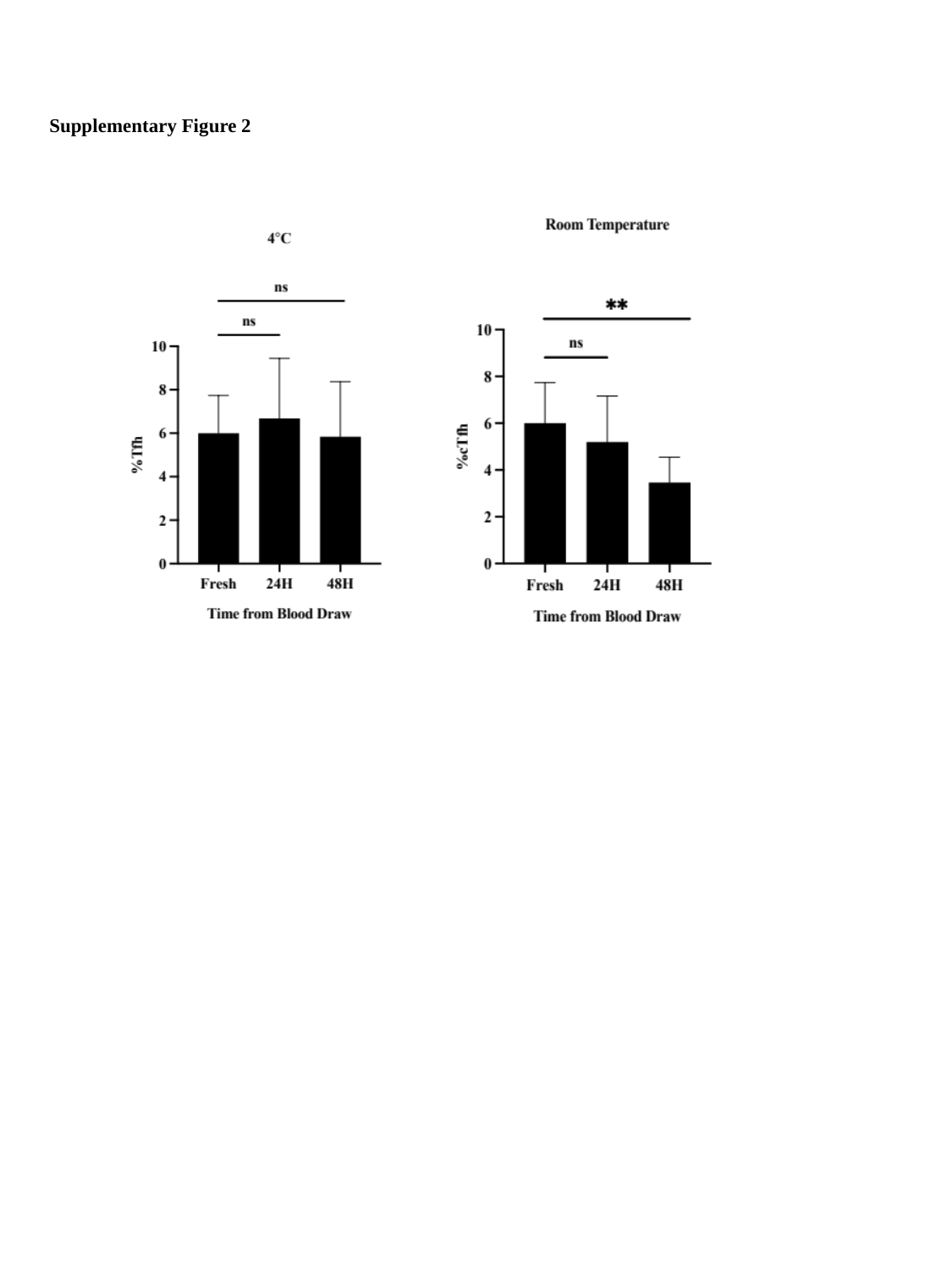

#### Supplementary Figure 2

### Slide 3
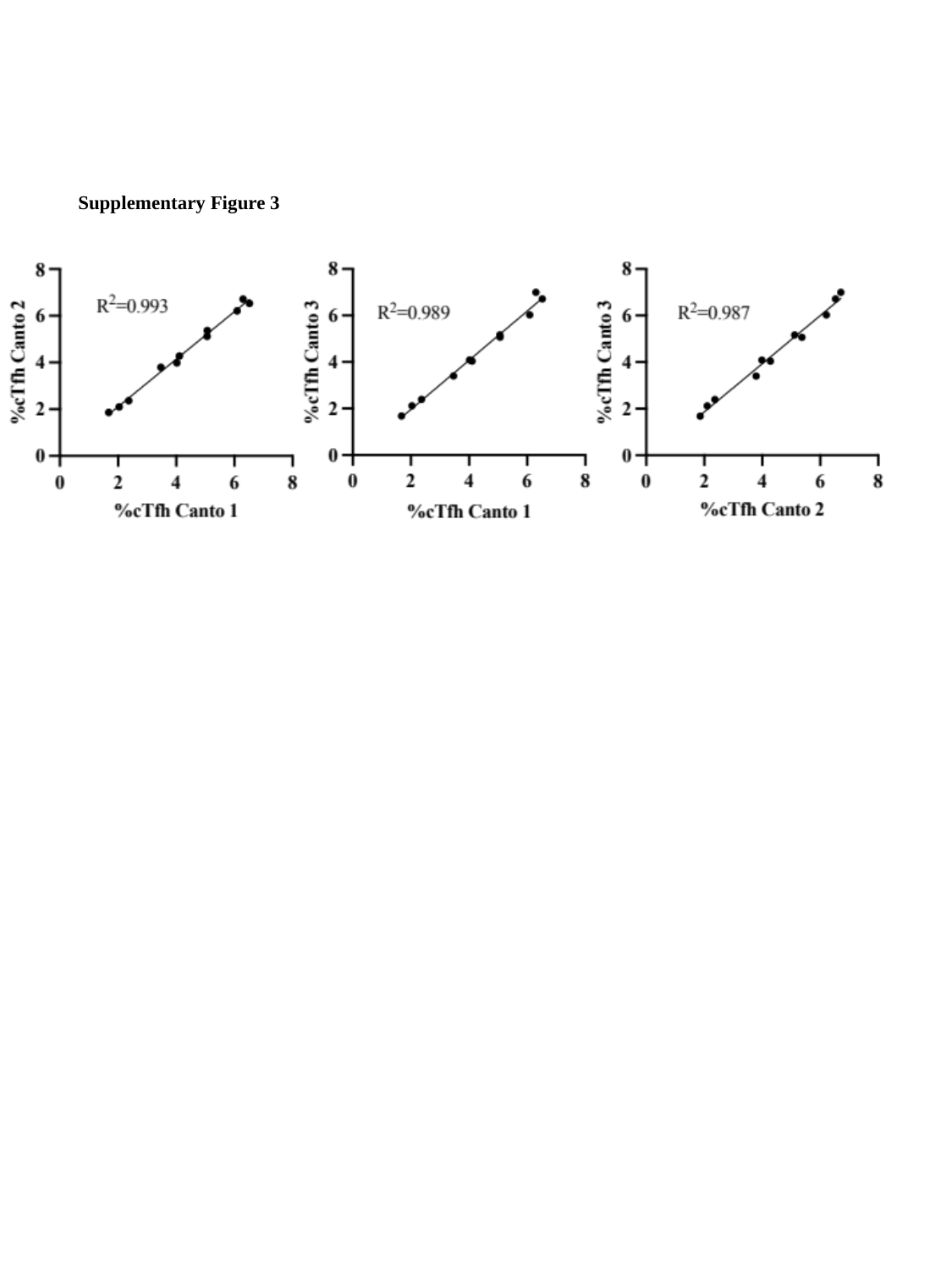

#### Supplementary Figure 3
